# Five-year changes in 24-hour sleep-wake activity and dementia risk in oldest old women

**DOI:** 10.1101/2024.07.23.24310882

**Authors:** Sasha Milton, Clémence Cavaillès, Sonia Ancoli-Israel, Katie L Stone, Kristine Yaffe, Yue Leng

## Abstract

**INTRODUCTION:** Sleep disruptions are associated with cognitive aging in older adults. However, it is unclear whether longitudinal changes in 24-hour multidimensional sleep-wake activity are linked to cognitive impairment in the oldest old.

**METHODS:** We studied 733 cognitively unimpaired women (mean age=82.5±2.9 years) who completed two actigraphy assessments over five years. We performed hierarchical clustering on principal components in nine sleep, napping, and circadian rest-activity rhythm parameters to identify multidimensional sleep-wake change profiles and multinomial logistic regression to evaluate the associations between sleep-wake changes and risk of cognitive impairment at follow-up.

**RESULTS:** We identified three sleep-wake change profiles: Stable Sleep (43.8%), Declining Nighttime Sleep (34.9%), and Increasing Sleepiness (21.3%). After adjustment for demographics and comorbidities, women with Increasing Sleepiness had approximately doubled (odds ratio=2.21, p=0.018) risk of dementia compared to those with Stable Sleep.

**DISCUSSION:** Increasing sleepiness may be an independent marker or risk factor for dementia in oldest old women.

## 1 INTRODUCTION

Sleep-wake disruption increases drastically in older adults with health comorbidities and is particularly common in older adults with Alzheimer’s disease and Alzheimer’s disease related dementias (AD/ADRD).[1,2] Growing evidence also suggests a potential bi-directional relationship between sleep and cognition, in which sleep disruption may not only stem from but also contribute to AD/ADRD pathology through the formation of amyloid plaques and tau pathology.[3,4] Indeed, epidemiological studies suggest that disrupted 24-hour sleep-wake activity, including extreme nighttime sleep duration, sleep fragmentation, excessive daytime napping, and disrupted circadian rest-activity rhythms (RARs), are associated with an increased risk of mild cognitive impairment (MCI) and dementia in late life.[5–9] Notably, a significant gap in existing research is the lack of longitudinal studies that follow participants over time to examine changes in sleep alongside changes in cognition. It remains unclear how 24-hour sleep-wake activity, including nighttime sleep, napping, and circadian RAR parameters, changes during the 8^th^ and 9^th^ decades of life. Furthermore, little is known about how these changes relate to dementia risk, particularly in the oldest old.

A few studies showed that declining or increasing self-reported sleep duration was associated with an increased risk of cognitive impairment.[10,11] However, self-reported measures of sleep are poorly correlated with objective measures and are prone to measurement errors, especially among older individuals.[12] Furthermore, sleep duration is only one aspect of sleep-wake activity; other dimensions such as nighttime sleep quality, daytime napping, and circadian disruption often exhibit more pronounced changes with advancing age,[13,14] and importantly, may be particularly involved in the neurodegenerative process.[15–18] Recent research has revealed an association between longitudinal changes in sleep stage percentages and decline in performance on cognitive tests in a cohort of older men.[19] However, it remains unclear whether longitudinal changes in objectively measured multidimensional 24-hour sleep-wake activity are associated with the risk of MCI and dementia in the oldest old, and which specific dimensions may be particularly relevant to cognitive aging. Given the significant shifts in sleep-wake patterns with age and the bi-directional link between sleep and cognitive aging, longitudinal change in 24-hour sleep-wake activity may be a useful marker or risk factor for cognitive impairment. Furthermore, multidimensional analyses of this change may capture information that assessments of sleep-wake parameters individually do not.

In a community-based cohort of oldest old women without cognitive impairment, we evaluated five-year changes in multidimensional 24-hour sleep-wake activity and their associations with the development of MCI and dementia over this period. First, we used a clustering analysis to characterize and identify multidimensional profiles of sleep-wake changes during the 8^th^ and 9^th^ decades of life. Next, we investigated the associations between these profiles of change and risk of developing MCI and dementia. Additionally, to determine which dimensions of sleep-wake change are most strongly associated with cognitive impairment, we also examined the change of each individual sleep-wake component separately. Finally, we included a sensitivity analysis in a subset of women who had apolipoprotein E (APOE) ε4 allele status information available in which we compared results with and without adjustment for APOEε4 status.

## 2 METHODS

### 2.1 Study design and participants

This longitudinal cohort study used data from the Study of Osteoporotic Fractures (SOF), in which community-dwelling women aged 65 years or older were enrolled between 1986 and 1988 from population-based listings in four United States areas: Baltimore, MD; Minneapolis, MN; Pittsburgh, PA; and Portland, OR. Study details have been previously described.[20] The institutional review boards at each site approved the study and all participants provided written informed consent. This analysis focused on 733 women without cognitive impairment at baseline who completed actigraphy at SOF clinic visits eight (2002-04, our study baseline) and nine (2006-08, follow-up visit) and had cognitive status evaluated at the follow-up visit. Figure 1 shows the full derivation of our sample.

**Figure 1.**
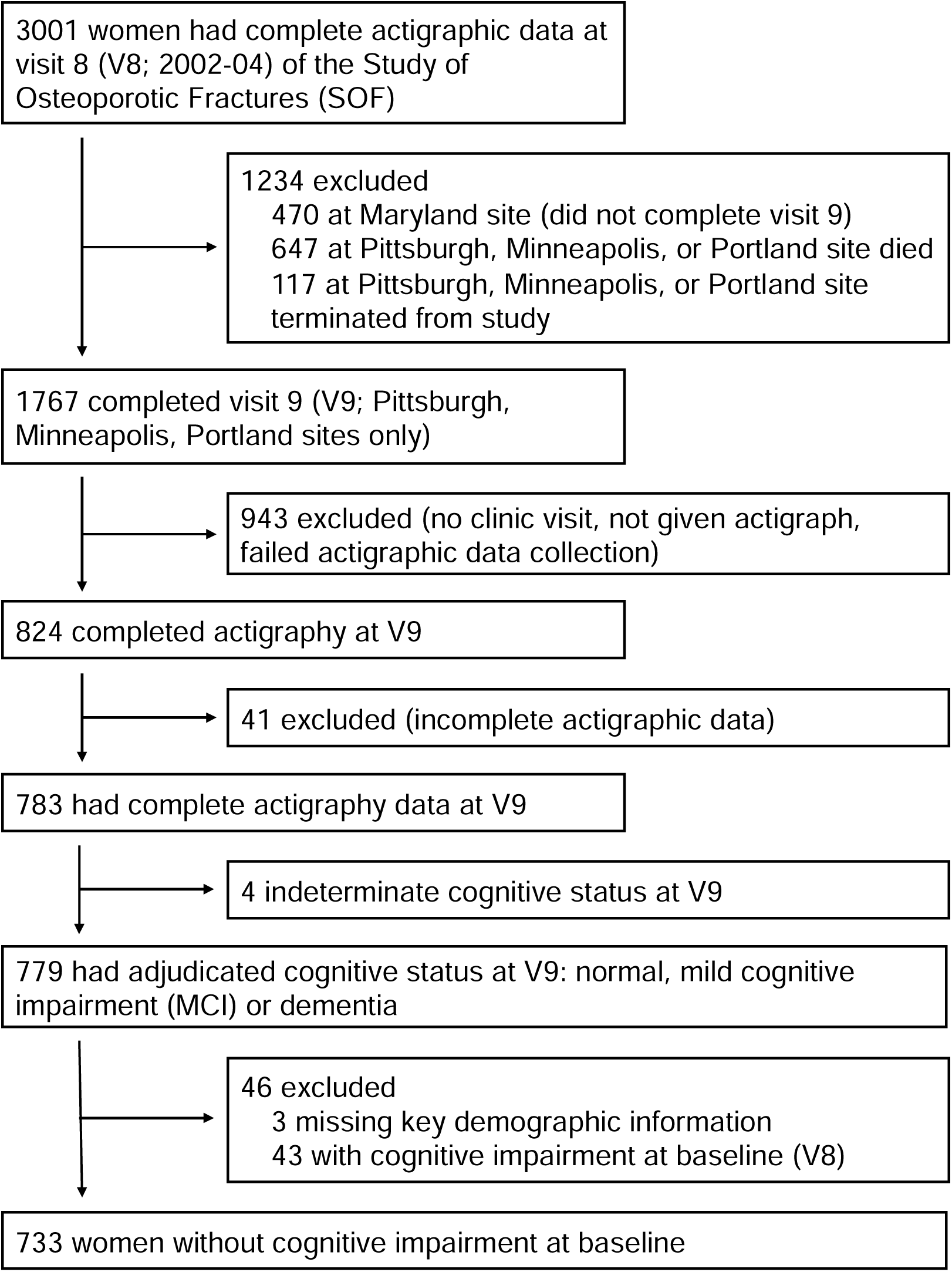
Derivation of the study sample from the Study of Osteoporotic Fractures.

### 2.2 Actigraphy and 24-hour sleep-wake parameters

We obtained objective measurement of 24-hour sleep-wake activity across a minimum of three consecutive days using wrist actigraphy (SleepWatch-O, Ambulatory Monitoring Inc, Ardsley, New York). Movements cause a piezoelectric bimorph-ceramic cantilevered balance beam to generate voltages, which are gathered continuously and summarized over one-minute intervals. The University of California, San Diego (UCSD) scoring algorithm in the Action W-2 software was used to differentiate sleep from wake times for each one-minute interval.[21] Proportional integration mode (PIM)[22] was used for data collection. Sleep logs were also completed by participants, allowing annotation and editing.

We investigated the five-year [median (range) = 5.0 (3.5,6.3) years] changes in nighttime sleep, napping, and circadian RAR parameters. Nighttime sleep parameters included total sleep time (TST or sleep duration; total minutes scored as sleep during the hours that participants reported being in bed during the night), sleep efficiency (SE; a measure of sleep quality calculated as the percentage of time in bed at night that was scored as sleep), and wake after sleep onset (WASO or sleep fragmentation; the number of minutes scored as wakefulness after sleep onset during the in-bed interval). According to the convention previously used in SOF, naps were defined as at least five consecutive minutes scored as sleep outside the main sleep interval.[23] Napping parameters included napping duration (total minutes scored as napping during daytime hours that participants reported not being in bed) and napping frequency (total number of nap episodes during daytime out-of-bed hours). For each participant, the mean value per day (across the full actigraphy recording) of each nighttime or napping parameter was calculated and used in analysis. Details of circadian RAR parameter calculations, which used an extended cosine model, have been described previously.[24] Circadian RAR parameters (calculated across the entire actigraphy recording) included (1) amplitude (counts/min), a measure of rhythm strength which is calculated as the difference in activity between the peak and the nadir at the point of greatest activity of the fitted curve; (2) mesor (counts/min), the mean activity level of the fitted curve; (3) robustness, which was assessed using a pseudo-F statistic representing goodness of extended cosine fit; and (4) acrophase (portions of hours), the timing of peak activity of the fitted curve. The five-year changes in all parameters were calculated as the follow-up value minus the baseline value, such that positive values indicated increases in parameters while negative values indicated decreases.

### 2.3 Main outcome

We excluded women with baseline probable cognitive impairment (defined as having a self-reported physician diagnosis of dementia, having a Mini-Mental State-Examination [MMSE] score <26, or having a Trail Making Test B [Trails B] completion time above 278 seconds).[25] Our primary outcome was cognitive status (normal cognition, MCI, or dementia) at the follow-up visit, which was determined using the following previously described process.[26] We used a screening based on (1) scores on cognitive tests, (2) self-reported physician diagnosis of dementia, and (3) residence in a nursing home. A woman who screened negative for all of these criteria was considered to be cognitively normal. If one or more of the criteria were met, the participant was included in a cognitive adjudication performed by a panel of clinical experts, using data from a neuropsychological battery performed at the follow-up visit and information on prior cognitive tests, demographics, medical history, medication use, symptoms of depression, and functional status. Diagnoses of MCI and dementia were based on criteria from a modified Peterson Criteria[27] and the Diagnostic and Statistical Manual of Mental Disorders (DSM-IV)[28], respectively. The remaining women who did not meet these criteria were considered to be cognitively normal.

### 2.4 Other variables

At study baseline, women completed a self-administered questionnaire that gathered information on age, education, race, walking for exercise (yes or no), and comorbidities (including self-reported diagnoses of diabetes, hypertension, stroke, and myocardial infarction). The Geriatric Depression Scale (GDS) was used to assess depressive symptoms, and the standard cutoff of ≥ 6 symptoms was used to define high depressive symptoms.[29] Women also completed a comprehensive examination and interview by a trained examiner. Body mass index (BMI) was calculated as weight in kilograms divided by the square of height in meters. Global cognition was assessed using the MMSE, which yields scores of 0-30, with higher scores indicating better cognition.[30] Participants also completed the Trails B, in which a faster completion time (in seconds) indicates better executive functioning.[31] Finally, participants were asked to show all current medications to examiners, allowing a complete inventory of current medication use to be recorded. Categorization of all medications was performed using a computerized medication coding dictionary.[32]

A subset of women from the Pittsburgh site also had serum collected at the SOF study baseline, allowing APOE phenotyping using isoelectric focusing and immunoblotting. APOEε4 status was collected as a binary variable (APOEε4 allele present vs. absent).[33]

### 2.5 Statistical analysis

We used hierarchical clustering on principal components (HCPC) to identify profiles of sleep-wake changes in women who had complete data for all nine 24-hour sleep and circadian RAR parameters at baseline and follow-up. We performed HCPC on the continuous five-year changes in the nine parameters using the “FactoMineR” and “factoextra” packages in R.[34] The number of principal components (PCs) necessary to explain 95% of cumulative variance were kept in the results of the principal components analysis (PCA) portion of the analysis. These PCs were used for unbiased hierarchical clustering in which the tree was cut at the suggested level (the partition point with the highest relative loss of inertia), revealing multiple distinct profiles of sleep-wake change. We first compared baseline characteristics by these profiles, using t-tests or ANOVA for continuous variables that were normally distributed, Kruskal-Wallis tests for skewed continuous data, and chi-square tests for categorical variables. Additionally, we divided the individual five-year sleep changes into two groups: “worsening” (women who showed a change of >1 standard deviation [SD] worse than the mean change), or “stable/improving”. For TST, SE, amplitude, mesor, and robustness, the “worsening” group represented significant decreases, while “worsening” represented significant increases for WASO and napping duration and frequency. We divided acrophase changes into an “earlier” group (women who showed a change of >1 SD below the mean, representing women whose peak activity moved earlier in the day) and a “stable/later” group.

To evaluate the associations between profiles of sleep-wake change (yielded from HCPC) and cognitive status, we used multinomial logistic regression models to calculate odds ratios (ORs) and 95% confidence intervals (CIs). In addition, we included separate models for each individual sleep or circadian RAR change parameter, with the “stable/improving” group (or the “stable/later” group, for acrophase) as a reference. We included unadjusted models in addition to models that adjusted for potential confounders including age, education, BMI, diabetes, hypertension, myocardial infarction, antidepressant use, and MMSE score at baseline. These were selected based on prior literature suggesting a plausible role in the relationship between sleep disruption and cognitive impairment. We also performed a sensitivity analysis in the subsample of women with APOEε4 data available and evaluated the impact of further adjustment for APOEε4 status on the association between sleep-wake change profiles/individual parameters and cognitive impairment. Statistical tests were 2-sided and used a p<.05 threshold. All analyses were performed in RStudio version 2023.12.0+369 (Posit Software, PBC).

## 3 RESULTS

The study sample included 733 women (9.3% Black) who were aged 82.5 ± 2.9 years at baseline. On the five-year follow-up, TST increased by 18.7 ± 73.3 min, SE increased by 0.94 ± 10.7%, WASO increased by 6.0 ± 45.0 min, napping duration increased by 33.1 ± 83.5 min, napping frequency increased by 1.8 ± 4.2 naps, acrophase moved earlier by 0.06 ± 1.0 hours, amplitude decreased by 675.9 ± 1074.4 counts/min, mesor decreased by 287.8 ± 553.4 counts/min, and robustness decreased by 190.4± 428.0 (pseudo-F). A total of six PCs explained 95.8% of the variance in the nine sleep and circadian RAR parameter changes included in the PCA, with PCs one, two, three, and four explaining 38.9%, 23.1%, 12.8%, and 8.9% of the variance, respectively. HCPC performed on these six PCs revealed three distinct profiles of change in parameters: Stable Sleep (SS; 321 women [43.8%]) was characterized by relative stability or small improvements; Declining Nighttime Sleep (DNS; 256 women [34.9%]) showed decreases in nighttime sleep quality and duration, moderate increases in napping parameters, and decreases in circadian RAR parameters; and Increasing Sleepiness (IS; 156 women [21.3%]) was characterized by large increases in both daytime and nighttime sleep duration and quality, in addition to decreases in circadian RAR parameters (Figure 2; Table 1).

**Figure 2.**
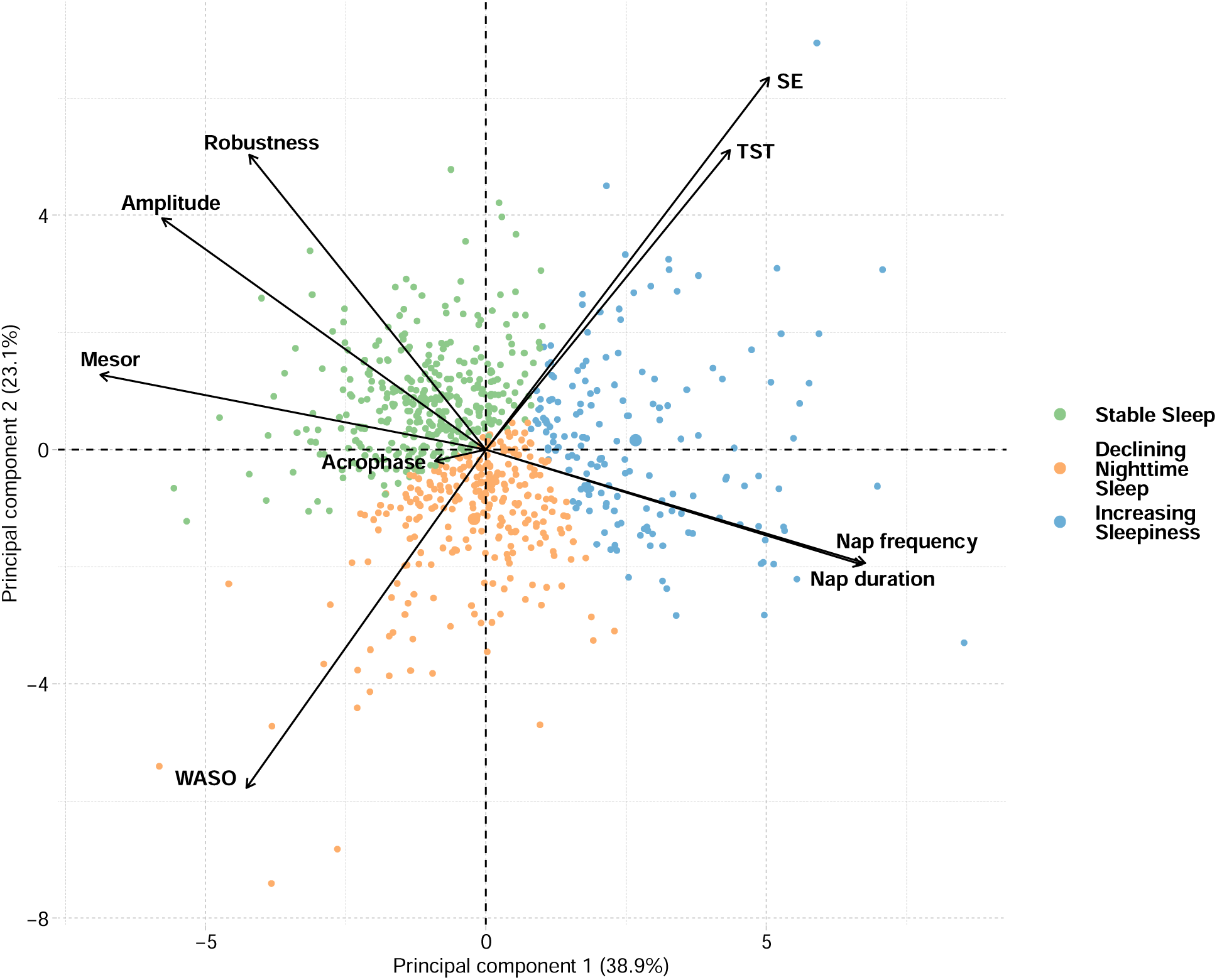
Principal components analysis biplot showing the three identified profiles of five-year changes in sleep and circadian rest-activity rhythm parameters. The black arrows show the loadings of the nine sleep or circadian rest-activity rhythm parameter changes with respect to principal components (PCs) 1 and 2, representing each parameter’s contribution to each PC. Small angles between arrows indicate high positive correlations between the variables. Participants on the same side as an arrow showed increases in that parameter while participants in the opposite direction showed decreases in that parameter. Abbreviations: TST, total sleep time; SE, sleep efficiency; WASO, wake after sleep onset.

**Table 1.** Profiles of five-year changes in 24-hour sleep-wake activity parameters.

| **Parameter (five-year change)** | **Stable sleep**<br>**(N=321)** | **Declining**<br>**Nighttime Sleep**<br>**(N=256)** | **Increasing**<br>**Sleepiness**<br>**(N=156)** | **P** |
| --- | --- | --- | --- | --- |
| **Total sleep time (min)** | 22.1 (68.8) | -14.7 (72.5) | 67.0 (86.8) | **<.001** |
| **Sleep efficiency (%)** | 0.87 (8.7) | -3.7 (8.5) | 9.0 (10.6) | **<.001** |
| **Wake after sleep onset (min)** | 3.6 (36.8) | 21.2 (38.6) | -23.8 (42.5) | **<.001** |
| **Napping duration (min)** | -7.7 (48.7) | 27.2 (60.0) | 104.4 (108.7) | **<.001** |
| **Napping frequency (count)** | -0.50 (2.8) | 1.3 (3.3) | 6.1 (7.0) | **<.001** |
| **Acrophase (portions of hours)** | -0.03 (0.95) | -0.09 (1.2) | -0.14 (1.3) | .12 |
| **Amplitude (counts/min)** | 4.8 (880.8) | -959.8 (824.7) | -1506.0 (1120.8) | **<.001** |
| **Mesor (counts/min)** | 0.66 (478.8) | -343.6 (393.7) | -804.6 (662.7) | **<.001** |
| **Robustness (pseudo-F)** | 72.0 (345.1) | -378.7 (399.5) | -398.3 (416.2) | **<.001** |

Note: Medians and interquartile ranges (IQRs) are reported.

Women with different profiles of sleep-wake changes had similar baseline characteristics (Table 2). With regards to baseline sleep and circadian RAR parameters, women exhibiting SS showed higher napping and weaker circadian RARs than women showing DNS or IS. Additionally, women with IS showed lower nighttime sleep quality and duration at baseline than women with SS or DNS (Supplementary Table 1). We also compared the changes in sleep-wake parameters across APOEε4 status and found that in comparison to those with no APOEε4 alleles, individuals with at least one APOEε4 allele showed worse changes in nighttime sleep parameters (decreases in TST and SE; increases in WASO) but better changes in napping (decreases) and circadian RAR (increases) parameters (Supplementary Table 2).

**Table 2.** Baseline characteristics by profiles of sleep-wake change.

|  | **Stable sleep**<br>**(N=321)** | **Declining**<br>**Nighttime Sleep**<br>**(N=256)** | **Increasing**<br>**Sleepiness**<br>**(N=156)** | **P** |
| --- | --- | --- | --- | --- |
| Age (years) | 82.4 (2.9) | 82.7 (2.8) | 82.5 (3.2) | .82 |
| <12 years education | 43 (13.4) | 36 (14.1) | 26 (16.7) | .63 |
| Black | 34 (10.6) | 14 (5.5) | 20 (12.8) | **.02** |
| Walks for exercise | 137 (42.7) | 108 (42.2) | 58 (37.2) | .54 |
| Body mass index (kg/m<sup>2</sup>) | 26.5 (5.5) | 26.8 (5.6) | 26.8 (6.8) | .55 |
| Diabetes | 35 (10.9) | 33 (12.9) | 16 (10.3) | .66 |
| Hypertension | 186 (57.9) | 157 (61.3) | 97 (62.2) | .59 |
| Stroke | 26 (8.1) | 26 (10.2) | 11 (7.1) | .50 |
| Myocardial infarction | 26 (8.1) | 23 (9.0) | 18 (11.5) | .47 |
| Depression | 16 (5.0) | 21 (8.2) | 17 (10.9) | .06 |
| MMSE score (0-30) | 28.8 (1.1) | 28.7 (1.1) | 28.6 (1.2) | .26 |
| Trails B score (0-180) | 123.0 (44.5) | 121.1 (44.1) | 126.4 (44.1) | .40 |
| Antidepressant user | 24 (7.5) | 33 (12.9) | 19 (12.2) | .07 |
| Sleep medication user | 55 (17.1) | 49 (19.1) | 28 (17.9) | .82 |

Note: Means and standard deviations are reported for age, MMSE score, and Trails B score. Median and interquartile range are reported for body mass index. Counts and percentages are reported for all other variables (categorical variables).
Abbreviations: MMSE, Mini-Mental State Examination.

During a follow-up time of five years [median (range) = 5.0 (3.5,6.3) years], 164 (22.4%) women developed MCI and 93 (12.7%) developed dementia. Women exhibiting DNS [OR (95% CI) = 2.03 (1.18,3.49)] or IS [OR (95% CI) = 2.85 (1.58,5.14)] had approximately two-fold to three-fold higher risk of dementia compared to women who showed SS. For individual parameters, women in the “worsening” group of the changes in SE, WASO, nap duration or nap frequency showed higher risk of dementia than women in the “stable/improving” group (Table 3).

**Table 3.** Unadjusted associations of changes in 24-hour sleep-wake activity parameters with MCI and dementia.

| **Parameter (five-year change) or profile** | **MCI (N=164)** | **Dementia (N=93)** |
| --- | --- | --- |
| **Profiles** |  |  |
| Stable sleep | 1.00 (ref) | 1.00 (ref) |
| Declining Nighttime Sleep | 0.83 (0.55,1.25) | **2.03 (1.18,3.49)** |
| Increasing Sleepiness | 1.21 (0.77,1.92) | **2.85 (1.58,5.14)** |
| **Total sleep time** |  |  |
| Stable/improving | 1.00 (ref) | 1.00 (ref) |
| Worsening | 0.83 (0.46,1.49) | 1.13 (0.58,2.21) |
| **Sleep efficiency** |  |  |
| Stable/improving | 1.00 (ref) | 1.00 (ref) |
| Worsening | 1.10 (0.59,2.04) | **2.44 (1.31,4.52)** |
| **Wake after sleep onset** |  |  |
| Stable/improving | 1.00 (ref) | 1.00 (ref) |
| Worsening | 1.34 (0.75,2.42) | **2.62 (1.42,4.80)** |
| **Nap duration** |  |  |
| Stable/improving | 1.00 (ref) | 1.00 (ref) |
| Worsening | 1.31 (0.76,2.26) | **2.44 (1.37,4.35)** |
| **Nap frequency** |  |  |
| Stable/improving | 1.00 (ref) | 1.00 (ref) |
| Worsening | 1.61 (0.96,2.68) | **2.49 (1.41,4.38)** |
| **Acrophase** |  |  |
| Stable/later | 1.00 (ref) | 1.00 (ref) |
| Earlier | 0.90 (0.49,1.65) | 0.96 (0.45,2.02) |
| **Amplitude** |  |  |
| Stable/improving | 1.00 (ref) | 1.00 (ref) |
| Worsening | 1.46 (0.87,2.46) | 1.77 (0.96,3.27) |
| **Mesor** |  |  |
| Stable/improving | 1.00 (ref) | 1.00 (ref) |
| Worsening | 1.41 (0.82,2.43) | 1.62 (0.85,3.08) |
| **Robustness** |  |  |
| Stable/improving | 1.00 (ref) | 1.00 (ref) |
| Worsening | 1.04 (0.61,1.77) | 1.70 (0.95,3.04) |

Note: Odds ratios and 95% confidence intervals are reported. Total N=733.
Abbreviations: MCI, mild cognitive impairment.

After adjustment for age, education, BMI, diabetes, hypertension, myocardial infarction, antidepressant use, and baseline cognition, women in the IS group had double risk of dementia [OR (95% CI) = 2.21 (1.14,4.26); p=0.018] compared to those in the SS group, whereas the association with dementia was attenuated for the DNS group [OR (95% CI) = 1.78 (0.98,3.26)]. When individual sleep-wake parameters were examined separately, the “worsening” group of changes in SE [OR (95% CI) = 2.15 (1.03,4.47)], WASO [OR (95% CI) = 2.26 (1.12,4.55)], nap duration [OR (95% CI) = 2.01 (1.03,3.93)], and nap frequency [OR (95% CI) = 2.19 (1.14,4.20)] were significantly associated with an approximately two-fold increased risk of dementia (Figure 3). TST and the circadian RAR parameters were not significantly associated with risk of dementia, and neither the profiles of sleep-wake change nor any of the individual parameters were associated with risk of MCI (Table 3; Figure 3). Finally, in a sensitivity analysis focused on 215 women who had APOEε4 status data available, results were similar with and without further adjustment for APOEε4 status (Supplementary Table 3).

**Figure 3.**
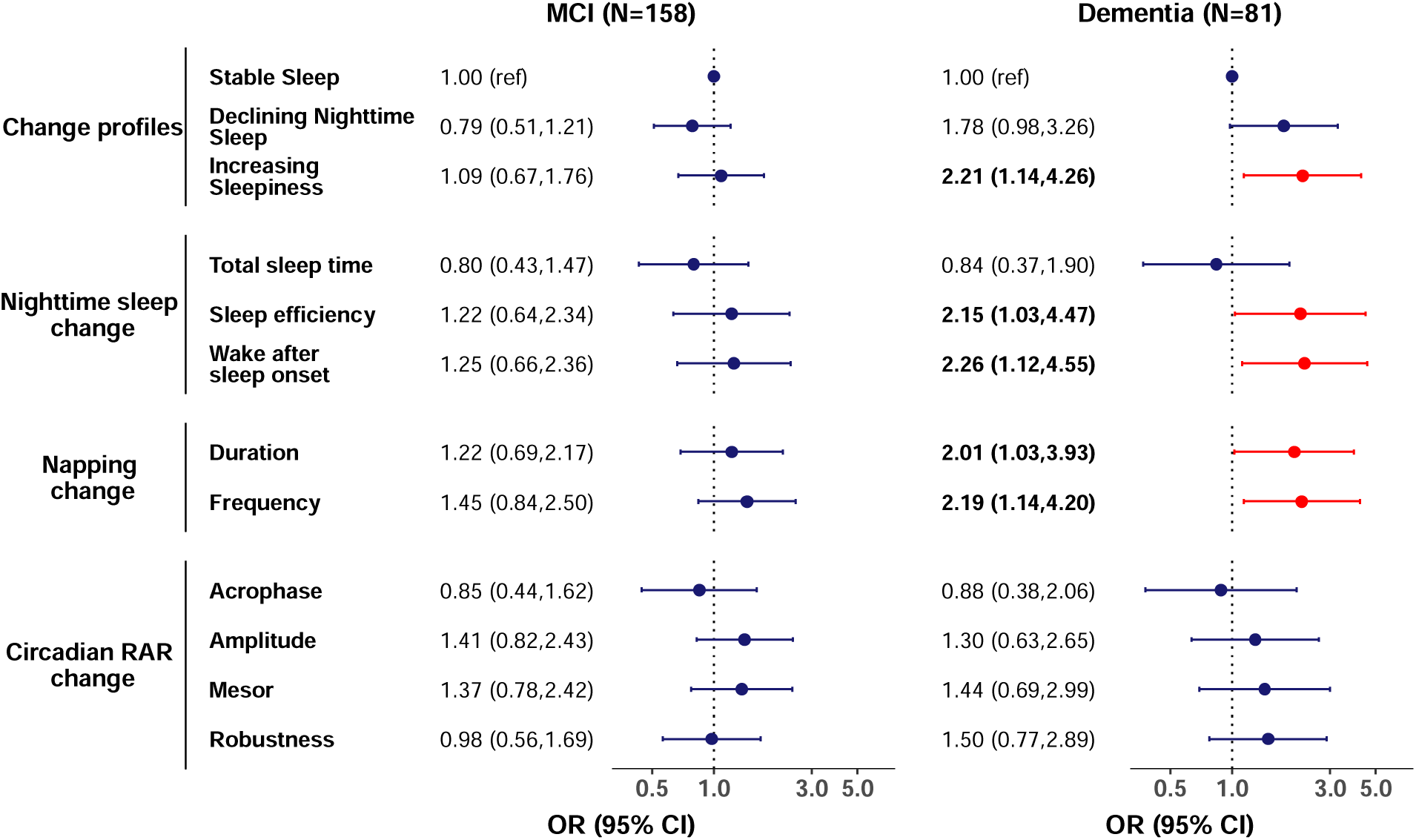
Multivariable-adjusted* associations of changes in sleep and circadian rest-activity rhythm parameters with MCI and dementia. Odds ratios (ORs) and 95% confidence intervals (95% CIs) are reported. Total N = 700. *Adjusted for age, race, education, body mass index, diabetes, hypertension, myocardial infarction, antidepressant use, and baseline cognition. The ORs and 95% CIs presented for individual parameters represent the odds of MCI/dementia for the “worsening” (“earlier” for acrophase) group in reference to the “stable/improving” (“stable/later” for acrophase) group. Abbreviations: MCI, mild cognitive impairment; RAR, rest-activity rhythm.

## 4 DISCUSSION

In a prospective cohort of oldest old women, we suggest that nighttime sleep, napping, and circadian RAR parameters can change drastically across only five years. We identified three profiles of five-year changes in multidimensional 24-hour sleep-wake activity: Stable Sleep (SS), Declining Nighttime Sleep (DNS), and Increasing Sleepiness (IS). Importantly, these profiles of change in multidimensional 24-hour sleep-wake activity were associated with development of cognitive impairment during this time. Compared to women whose sleep remained stable, those showing large increases in 24-hour sleepiness were approximately twice as likely to develop dementia, but not MCI, independent of other factors, including demographics, comorbidities, and APOEε4 status. These data suggest, for the first time, that longitudinal change in multidimensional 24-hour sleep-wake activity, particularly increasing sleepiness, may be a marker of dementia risk in oldest old women.

Our study extends a mounting body of research on sleep disturbances and risk of ADRD [5,6,25,35–37] by highlighting the importance of changes in multidimensional 24-hour sleep-wake activity over time in the oldest old. This adds to prior research suggesting relatively stable sleep duration trajectories from the 40s to the 70s, with no observed links to cognition or brain structure in late life.[38] In our study of oldest old women, less than half had Stable Sleep over five years. One-third showed large decreases in nighttime sleep quality and duration (i.e., Declining Nighttime Sleep), and 21% experienced large increases in both daytime and nighttime sleep duration and quality (i.e., Increasing Sleepiness). While it is surprising to note such marked changes in sleep-wake activity in a span of only five years among women in their 80s, these changes may represent important missing pieces of the puzzle on sleep and AD/ADRD.

Indeed, older women with Increasing Sleepiness exhibited a doubled risk of developing dementia, even after accounting for multiple comorbidities, while the association for Declining Nighttime Sleep was attenuated after adjustment for covariates. This corroborates recent evidence suggesting a bi-directional relationship between daytime napping and cognitive decline.[18] The elevated risk of dementia among participants with both increasing daytime and nighttime sleep duration and efficiency also aligns with prior findings that indicate an association between napping and cognitive impairment only among those with higher sleep efficiency and adequate sleep duration.[6] Interestingly, while the high-risk Increasing Sleepiness group showed increases in sleep efficiency and decreases in WASO, we found that when analyzed individually, changes in the opposite direction (decreases in sleep efficiency and increases in WASO) were associated with increased dementia risk. Comprehensive examinations of 24-hour sleep-wake activity can thus reveal a high risk of dementia in individuals that may be overlooked in analyses of individual parameters. Indeed, there has been increasing interest in viewing sleep health as a multidimensional concept.[39–41] One recent study found that older men with a higher number of poor subjective sleep health dimensions had greater cognitive decline over ten years.[42] Notably, our findings conflict with another study that identified sleep health profiles measured by actigraphy at a single timepoint, which showed a significantly increased mortality risk associated only with the ‘Inadequate Sleep’ profile, but not the ‘High Sleep Propensity’ profile (high napping and high maintenance; similar to our IS group).[43] It is possible that longitudinal changes, rather than one-time assessment of 24-hour sleepiness, may have specific health implications, or that these implications may be particular to cognitive impairment rather than other health risks. The current study is the first, to our knowledge, to suggest that the co-occurrence of increasing daytime and nighttime sleep over time may signal an underlying dementia risk in oldest old women.

When individual sleep-wake change parameters were examined separately, the magnitudes of the associations we observed for changes in nighttime sleep efficiency, WASO, and napping parameters were stronger than what has been previously reported for one-time assessments of these parameters in the same cohort.[5,25,35] In contrast to previous research that found an association between circadian RAR parameters and risk of dementia, [5] we did not find a significant association between longitudinal changes in circadian RAR parameters and dementia risk, despite similar effect sizes. This may be due to the limited number of women who had 24-hour sleep-wake activity assessed at multiple timepoints, resulting in lower statistical power compared to the prior study. Additionally, several studies reported links between changes in self-reported sleep duration and cognitive impairment in middle-aged and older adults.[10,11,44] The current study, however, did not find an association between change in objective sleep duration alone and risk of cognitive impairment. These divergent findings may result from differences in self-report and objective measures. It has been posited that the quality of sleep, rather than its quantity, may hold greater importance for cognitive health.[45] Moreover, given the multitude of factors that can affect sleep duration over five years in the elderly, such changes may not serve as sensitive indicators of cognitive decline. The current findings emphasize the need to use objective measures to examine all aspects of 24-hour sleep-wake activity and at multiple time points.

Changes in 24-hour sleep-wake activity can occur in the normal aging process and can be exacerbated by AD/ADRD pathology in the preclinical stage.[1,17,46] Sleep-wake instability, including decreased sleep duration and increased sleep fragmentation, may increase with normal and pathological aging due to structural changes in wake-promoting nuclei in the lateral hypothalamic area and the locus coeruleus, as well as in sleep-promoting nuclei in the preoptic area.[1] Evidence from both animal and human studies suggests that amyloid-β and tau pathology (two hallmarks of AD/ADRD that can begin to accumulate several decades prior to cognitive impairment[47]) could affect these sleep-wake centers, thereby leading to deterioration of nighttime sleep quality over the years preceding clinical AD/ADRD.[48] Postmortem evidence has revealed a loss of wake-promoting neurons in AD patients, which may help explain the elevated risk of dementia in the IS group.[16,49] Meanwhile, the possibility of a bi-directional relationship in which sleep changes themselves play a role in AD/ADRD pathogenesis should not be overlooked. Evidence suggests that disrupted 24-hour sleep-wake activity can result in AD/ADRD pathology, possibly due to increased amyloid-β plaque formation and tau deposition, in addition to ineffective clearance via the glymphatic system.[51–54] Therefore, concurrent declines in sleep health and cognition may exacerbate one another, creating a vicious cycle and accelerating deterioration in both domains. Notably, we did not observe an association for sleep change and MCI, which may be due to the high heterogeneity and mixed etiologies of MCI.[55] Further studies are needed to elucidate the pathophysiological pathways underlying changes in multidimensional 24-hour sleep-wake activity across the AD/ADRD spectrum.

A major strength of this study is the objective and repeated assessment of multidimensional 24-hour sleep-wake activity, including nighttime sleep, napping, and circadian RAR measures, allowing for a comprehensive evaluation of change in 24-hour sleep-wake activity in the oldest old. This is a first in research of sleep and AD/ADRD. Additionally, classifications of MCI and dementia at follow-up were determined through a neuropsychological battery and adjudication process. There are a few limitations. First, this study included mostly white women in their 80s, and thus our findings may not be generalizable to men or more diverse populations. Additionally, sleep-wake change was evaluated over a period of only five years; cognitive status was evaluated at the same timepoint as the second sleep evaluation, and thus the development of dementia may have occurred in parallel to the change in sleep. However, we have used stringent criteria to exclude women with any evidence of cognitive impairment at baseline; additionally, those with different profiles of sleep-wake change had similar baseline characteristics, including cognition. Future studies with multiple assessments of sleep earlier in life and with longer follow-up for incident dementia after the measurement of sleep-wake change will help clarify the temporal relationship. As AD biomarkers were not assessed in this study, our ability to draw conclusions concerning the relationship between sleep-wake changes and AD pathology was limited, though our consideration of APOEε4 status was a strength. Relative to individuals without the APOEε4 allele, those with at least one APOEε4 allele showed five-year decreases in napping and improvements in circadian RAR parameters, suggesting that the observed changes in these parameters may not only be markers of cognitive impairment but may be early risk factors. Ultimately, due to the observational nature of our study, causal inferences cannot be derived. Randomized controlled trials are needed to determine whether improvement of 24-hour sleep-wake activity may help slow down cognitive decline.

### 4.1 Conclusion

Sleep-wake activity can change drastically in oldest old women. Among community-dwelling women in their 80s, those with increasing 24-hour sleepiness (large increases in both daytime and nighttime sleep) over five years had double the risk of dementia during the same time period. Changes in multidimensional 24-hour sleep-wake activity may be intertwined with cognitive aging and may serve as an early marker or risk factor for dementia in the oldest old. Our findings emphasize the importance of a holistic view of multiple dimensions of 24-hour sleep-wake activity and the need for research on the link between longitudinal sleep-wake change and dementia risk.

## Data Availability

All data produced in the present study are available upon reasonable request to the authors.

https://sofonline.ucsf.edu/

## Acknowledgements

No acknowledgements are included.

## Funding/support and role of funder/supporter

This study was supported by the National Institutes of Health (NIH) National Institute on Aging grants R21AG085495 and R01AG083836 (Dr. Leng), and 5R01AG026720 (Drs. Yaffe and Stone). The Study of Osteoporotic Fractures (SOF) was supported by NIH grants (AG021918, AG026720, AG05394, AG05407, AG08415, AR35582, AR35583, AR35584, RO1 AG005407, R01 AG027576-22, 2 R01 AG005394-22A1, 2 RO1 AG027574-22A1, HL40489, T32 AG000212-14). The funding sources had no role in the design and conduct of the study; collection, management, analysis, and interpretation of the data; preparation, review, or approval of the manuscript; and decision to submit the manuscript for publication.

## REFERENCES

[1] Mander BA, Winer JR, Walker MP. Sleep and Human Aging. Neuron 2017;94:19–36. 10.1016/j.neuron.2017.02.004.

[2] Porter VR, Buxton WG, Avidan AY. Sleep, Cognition and Dementia. Curr Psychiatry Rep 2015;17:97. 10.1007/s11920-015-0631-8.

[3] Ju Y-ES, Lucey BP, Holtzman DM. Sleep and Alzheimer disease pathology—a bidirectional relationship. Nat Rev Neurol 2014;10:115–9. 10.1038/nrneurol.2013.269.

[4] Lucey BP. It’s complicated: The relationship between sleep and Alzheimer’s disease in humans. Neurobiol Dis 2020;144:105031. 10.1016/j.nbd.2020.105031.

[5] Tranah GJ, Blackwell T, Stone KL, Ancoli Israel S, Paudel ML, Ensrud KE, et al. Circadian activity rhythms and risk of incident dementia and mild cognitive impairment in older women. Ann Neurol 2011;70:722–32. 10.1002/ana.22468.

[6] Leng Y, Redline S, Stone KL, Ancoli Israel S, Yaffe K. Objective napping, cognitive decline, and risk of cognitive impairment in older men. Alzheimer’s & Dementia 2019;15:1039–47. 10.1016/j.jalz.2019.04.009.

[7] Cox SR, Ritchie SJ, Allerhand M, Hagenaars SP, Radakovic R, Breen DP, et al. Sleep and cognitive aging in the eighth decade of life. Sleep 2019;42. 10.1093/sleep/zsz019.

[8] Li P, Gao L, Gaba A, Yu L, Cui L, Fan W, et al. Circadian disturbances in Alzheimer’s disease progression: a prospective observational cohort study of community-based older adults. Lancet Healthy Longev 2020;1:e96–105. 10.1016/S2666-7568(20)30015-5.

[9] Posner AB, Tranah GJ, Blackwell T, Yaffe K, Ancoli-Israel S, Redline S, et al. Predicting incident dementia and mild cognitive impairment in older women with nonparametric analysis of circadian activity rhythms in the Study of Osteoporotic Fractures. Sleep 2021;44. 10.1093/sleep/zsab119.

[10] Keil SA, Schindler AG, Wang MX, Piantino J, Silbert LC, Elliott JE, et al. Longitudinal Sleep Patterns and Cognitive Impairment in Older Adults. JAMA Netw Open 2023;6:e2346006. 10.1001/jamanetworkopen.2023.46006.

[11] Sabia S, Fayosse A, Dumurgier J, van Hees VT, Paquet C, Sommerlad A, et al. Association of sleep duration in middle and old age with incidence of dementia. Nat Commun 2021;12:2289. 10.1038/s41467-021-22354-2.

[12] Miner B, Stone KL, Zeitzer JM, Han L, Doyle M, Blackwell T, et al. Self-reported and actigraphic short sleep duration in older adults. Journal of Clinical Sleep Medicine 2022;18:403–13. 10.5664/jcsm.9584.

[13] Taillard J, Gronfier C, Bioulac S, Philip P, Sagaspe P. Sleep in Normal Aging, Homeostatic and Circadian Regulation and Vulnerability to Sleep Deprivation. Brain Sci 2021;11:1003. 10.3390/brainsci11081003.

[14] Duffy JF, Zitting K-M, Chinoy ED. Aging and Circadian Rhythms. Sleep Med Clin 2015;10:423–34. 10.1016/j.jsmc.2015.08.002.

[15] Leng Y, Musiek ES, Hu K, Cappuccio FP, Yaffe K. Association between circadian rhythms and neurodegenerative diseases. Lancet Neurol 2019;18:307–18. 10.1016/S1474-4422(18)30461-7.

[16] Oh JY, Walsh CM, Ranasinghe K, Mladinov M, Pereira FL, Petersen C, et al. Subcortical Neuronal Correlates of Sleep in Neurodegenerative Diseases. JAMA Neurol 2022;79:498. 10.1001/jamaneurol.2022.0429.

[17] Musiek ES, Bhimasani M, Zangrilli MA, Morris JC, Holtzman DM, Ju Y-ES. Circadian Rest-Activity Pattern Changes in Aging and Preclinical Alzheimer Disease. JAMA Neurol 2018;75:582. 10.1001/jamaneurol.2017.4719.

[18] Li P, Gao L, Yu L, Zheng X, Ulsa MC, Yang H, et al. Daytime napping and Alzheimer’s dementia: A potential bidirectional relationship. Alzheimer’s & Dementia 2023;19:158–68. 10.1002/alz.12636.

[19] Wang Q, Stone KL, Lu Z, Tian S, Zheng Y, Zhao B, et al. Associations between Longitudinal Changes in Sleep Stages and Risk of Cognitive Decline in Older Men. Sleep 2024. 10.1093/sleep/zsae125.

[20] Cummings SR, Browner W, Black DM, Nevitt MC, Browner W, Genant HK, et al. Bone density at various sites for prediction of hip fractures. The Lancet 1993;341:72–5. 10.1016/0140-6736(93)92555-8.

[21] Jean-Louis G, Kripke DF, Mason WJ, Elliott JA, Youngstedt SD. Sleep estimation from wrist movement quantified by different actigraphic modalities. J Neurosci Methods 2001;105:185–91. 10.1016/S0165-0270(00)00364-2.

[22] Blackwell T, Redline S, Ancoli-Israel S, Schneider JL, Surovec S, Johnson NL, et al. Comparison of Sleep Parameters from Actigraphy and Polysomnography in Older Women: The SOF Study. Sleep 2008;31:283–91. 10.1093/sleep/31.2.283.

[23] Leng Y, Stone K, Ancoli-Israel S, Covinsky K, Yaffe K. Who Take Naps? Self-Reported and Objectively Measured Napping in Very Old Women. The Journals of Gerontology: Series A 2018;73:374–9. 10.1093/gerona/glx014.

[24] Marler MR, Gehrman P, Martin JL, Ancoli Israel S. The sigmoidally transformed cosine curve: a mathematical model for circadian rhythms with symmetric non sinusoidal shapes. Stat Med 2006;25:3893–904. 10.1002/sim.2466.

[25] Blackwell T, Yaffe K, Ancoli-Israel S, Schneider JL, Cauley JA, Hillier TA, et al. Poor Sleep Is Associated With Impaired Cognitive Function in Older Women: The Study of Osteoporotic Fractures. J Gerontol A Biol Sci Med Sci 2006;61:405–10. 10.1093/gerona/61.4.405.

[26] Yaffe K, Middleton LE, Lui L-Y, Spira AP, Stone K, Racine C, et al. Mild Cognitive Impairment, Dementia, and Their Subtypes in Oldest Old Women. Arch Neurol 2011;68. 10.1001/archneurol.2011.82.

[27] Petersen RC, Doody R, Kurz A, Mohs RC, Morris JC, Rabins P V., et al. Current Concepts in Mild Cognitive Impairment. Arch Neurol 2001;58:1985. 10.1001/archneur.58.12.1985.

[28] Guze SB. Diagnostic and Statistical Manual of Mental Disorders, 4th ed. (DSM-IV). American Journal of Psychiatry 1995;152:1228–1228. 10.1176/ajp.152.8.1228.

[29] Yesavage JA, Sheikh JI. 9/Geriatric Depression Scale (GDS). Clin Gerontol 1986;5:165– 73. 10.1300/J018v05n01_09.

[30] Folstein MF. The Mini-Mental State Examination. Arch Gen Psychiatry 1983;40:812. 10.1001/archpsyc.1983.01790060110016.

[31] Reitan RM. VALIDITY OF THE TRAIL MAKING TEST AS AN INDICATOR OF ORGANIC BRAIN DAMAGE. Percept Mot Skills 1958;8:271. 10.2466/PMS.8.7.271-276.

[32] Pahor M, Chrischilles EA, Guralnik JM, Brown SL, Wallace RB, Carbonin P. Drug data coding and analysis in epidemiologic studies. Eur J Epidemiol 1994;10:405–11. 10.1007/BF01719664.

[33] Cauley JA, Zmuda JM, Yaffe K, Kuller LH, Ferrell RE, Wisniewski SR, et al. Apolipoprotein E Polymorphism: A New Genetic Marker of Hip Fracture RiskThe Study of Osteoporotic Fractures. Journal of Bone and Mineral Research 1999;14:1175–81. 10.1359/jbmr.1999.14.7.1175.

[34] Husson F, Le S, Pagès J. Exploratory Multivariate Analysis by Example Using R. Chapman and Hall/CRC; 2017. 10.1201/b21874.

[35] Diem SJ, Blackwell TL, Stone KL, Yaffe K, Tranah G, Cauley JA, et al. Measures of Sleep–Wake Patterns and Risk of Mild Cognitive Impairment or Dementia in Older Women. The American Journal of Geriatric Psychiatry 2016;24:248–58. 10.1016/j.jagp.2015.12.002.

[36] Lu Z, Leung JCS, Feng H, Zhang J, Wing YK, Kwok TCY. Circadian rest-activity rhythms and cognitive decline and impairment in older Chinese adults: A multicohort study with prospective follow-up. Arch Gerontol Geriatr 2024;116:105215. 10.1016/j.archger.2023.105215.

[37] Xiao Q, Shadyab AH, Rapp SR, Stone KL, Yaffe K, Sampson JN, et al. Rest activity rhythms and cognitive impairment and dementia in older women: Results from the Women’s Health Initiative. J Am Geriatr Soc 2022;70:2925–37. 10.1111/jgs.17926.

[38] Zitser J, Anatürk M, Zsoldos E, Mahmood A, Filippini N, Suri S, et al. Sleep duration over 28 years, cognition, gray matter volume, and white matter microstructure: a prospective cohort study. Sleep 2020;43. 10.1093/sleep/zsz290.

[39] Buysse DJ. Sleep Health: Can We Define It? Does It Matter? Sleep 2014;37:9–17. 10.5665/sleep.3298.

[40] Wallace ML, Yu L, Buysse DJ, Stone KL, Redline S, Smagula SF, et al. Multidimensional sleep health domains in older men and women: an actigraphy factor analysis. Sleep 2021;44. 10.1093/sleep/zsaa181.

[41] Wallace ML, Buysse DJ, Redline S, Stone KL, Ensrud K, Leng Y, et al. Multidimensional Sleep and Mortality in Older Adults: A Machine-Learning Comparison With Other Risk Factors. The Journals of Gerontology: Series A 2019;74:1903–9. 10.1093/gerona/glz044.

[42] Cavaillès C, Yaffe K, Blackwell T, Buysse D, Stone K, Leng Y. Multidimensional Sleep Health and Long-Term Cognitive Decline in Community-Dwelling Older Men. Journal of Alzheimer’s Disease 2023;96:65–71. 10.3233/JAD-230737.

[43] Wallace ML, Lee S, Stone KL, Hall MH, Smagula SF, Redline S, et al. Actigraphy-derived sleep health profiles and mortality in older men and women. Sleep 2022;45. 10.1093/sleep/zsac015.

[44] Zhu Q, Fan H, Zhang X, Ji C, Xia Y. Changes in sleep duration and 3-year risk of mild cognitive impairment in Chinese older adults. Aging 2020;12:309–17. 10.18632/aging.102616.

[45] Leng Y, Knutson K, Carnethon MR, Yaffe K. Association Between Sleep Quantity and Quality in Early Adulthood With Cognitive Function in Midlife. Neurology 2024;102. 10.1212/WNL.0000000000208056.

[46] Ju Y-ES, McLeland JS, Toedebusch CD, Xiong C, Fagan AM, Duntley SP, et al. Sleep Quality and Preclinical Alzheimer Disease. JAMA Neurol 2013;70:587. 10.1001/jamaneurol.2013.2334.

[47] Jack CR, Knopman DS, Jagust WJ, Shaw LM, Aisen PS, Weiner MW, et al. Hypothetical model of dynamic biomarkers of the Alzheimer’s pathological cascade. Lancet Neurol 2010;9:119–28. 10.1016/S1474-4422(09)70299-6.

[48] Wang C, Holtzman DM. Bidirectional relationship between sleep and Alzheimer’s disease: role of amyloid, tau, and other factors. Neuropsychopharmacology 2020;45:104–20. 10.1038/s41386-019-0478-5.

[49] Oh J, Eser RA, Ehrenberg AJ, Morales D, Petersen C, Kudlacek J, et al. Profound degeneration of wake promoting neurons in Alzheimer’s disease. Alzheimer’s & Dementia 2019;15:1253–63. 10.1016/j.jalz.2019.06.3916.

[50] Wang JL, Lim AS, Chiang W, Hsieh W, Lo M, Schneider JA, et al. Suprachiasmatic neuron numbers and rest–activity circadian rhythms in older humans. Ann Neurol 2015;78:317–22. 10.1002/ana.24432.

[51] Musiek ES, Holtzman DM. Mechanisms linking circadian clocks, sleep, and neurodegeneration. Science (1979) 2016;354:1004–8. 10.1126/science.aah4968.

[52] Carvalho DZ, St Louis EK, Knopman DS, Boeve BF, Lowe VJ, Roberts RO, et al. Association of Excessive Daytime Sleepiness With Longitudinal β-Amyloid Accumulation in Elderly Persons Without Dementia. JAMA Neurol 2018;75:672. 10.1001/jamaneurol.2018.0049.

[53] Spira AP, An Y, Wu MN, Owusu JT, Simonsick EM, Bilgel M, et al. Excessive daytime sleepiness and napping in cognitively normal adults: associations with subsequent amyloid deposition measured by PiB PET. Sleep 2018;41. 10.1093/sleep/zsy152.

[54] Holth JK, Fritschi SK, Wang C, Pedersen NP, Cirrito JR, Mahan TE, et al. The sleep-wake cycle regulates brain interstitial fluid tau in mice and CSF tau in humans. Science (1979) 2019;363:880–4. 10.1126/science.aav2546.

[55] Abner EL, Kryscio RJ, Schmitt FA, Fardo DW, Moga DC, Ighodaro ET, et al. Outcomes after diagnosis of mild cognitive impairment in a large autopsy series. Ann Neurol 2017;81:549–59. 10.1002/ana.24903.

