## Supplementary Materials for "Five-year changes in 24-hour sleep-wake activity and dementia risk in oldest old women"

| Supplementary Table 1. Baseline sleep, napping and circadian rest-activity rhythm parameters by profiles of sleep-wake change | | | | |
| --- | --- | --- | --- | --- |
| Parameter | **Stable Sleep**  **(N=321)** | **Declining**  **Nighttime Sleep**  **(N=256)** | **Increasing**  **Sleepiness**  **(N=156)** | **P** |
| Total sleep time (min) | 413.0 (82.8) | 417.1 (72.9) | 405.4 (89.3) | **.045** |
| Sleep efficiency (%) | 82.8 (11.0) | 82.9 (10.7) | 78.3 (14.0) | **<.001** |
| Wake after sleep onset (min) | 52.0 (45.0) | 54.0 (39.2) | 71.0 (63.3) | **<.001** |
| Napping duration (min) | 67.3 (71.3) | 40.5 (54.5) | 47.0 (51.9) | **<.001** |
| Napping frequency (count) | 3.5 (3.7) | 2.3 (3.2) | 2.3 (3.0) | **<.001** |
| Acrophase (portions of hours) | 14.8 (1.4) | 14.8 (1.3) | 14.7 (1.2) | .74 |
| Amplitude (counts/min) | 3432.9 (1082.7) | 3903.4 (1056.9) | 3661.0 (1125.6) | **<.001** |
| Mesor (counts/min) | 2078.0 (577.8) | 2260.1 (541.2) | 2227.3 (594.7) | **<.001** |
| Robustness (pseudo-F) | 760.6 (464.6) | 1029.1 (610.4) | 919.7 (508.3) | **<.001** |
| Note: Medians and interquartile ranges (IQRs) are reported. | | | | |

| Supplementary Table 2. Changes in sleep, napping and circadian rest-activity rhythm parameters by APOEε4 status | | | |
| --- | --- | --- | --- |
| Parameter (five-year change) | **APOEε4 allele absent**  **(N=198)** | **APOEε4 allele present**  **(N=17)** | **P** |
| Total sleep time (min) | 7.2 (87.8) | -26.8 (63.5) | 0.09 |
| Sleep efficiency (%) | -0.03 (9.4) | -4.0 (4.5) | **0.01** |
| Wake after sleep onset (min) | 11.1 (40.1) | 18.1 (42.2) | 0.07 |
| Napping duration (min) | 20.4 (80.4) | -13.7 (43.4) | **0.01** |
| Napping frequency (count) | 1.2 (4.7) | -0.33 (2.2) | **0.03** |
| Acrophase (portions of hours) | -0.24 (1.0) | -0.08 (0.97) | 0.35 |
| Amplitude (counts/min) | -727.7 (1487.7) | 133.4 (1481.5) | **0.02** |
| Mesor (counts/min) | -250.8 (671.2) | 33.4 (584.1) | 0.08 |
| Robustness (pseudo-F) | -238.2 (530.6) | -103.1 (572.1) | 0.28 |
| Note: Medians and interquartile ranges (IQRs) are reported. | | | |

| **Supplementary Table 3. Associations of changes in 24-hour sleep-wake activity with MCI and dementia with and without adjustment* for APOEε4 status** | | | | |
| --- | --- | --- | --- | --- |
| **Parameter (five-year change) or profile** | **Not adjusted for APOEε4 status** | | **Adjusted for APOEε4 status** | |
|  | **MCI (N=45)** | **Dementia (N=25)** | **MCI (N=45)** | **Dementia (N=25)** |
| **Profiles** |  |  |  |  |
| Stable sleep | 1.00 (ref) | 1.00 (ref) | 1.00 (ref) | 1.00 (ref) |
| Declining Nighttime Sleep | 0.49 (0.22,1.09) | 2.10 (0.70,6.30) | 0.49 (0.22,1.09) | 2.36 (0.75,7.41) |
| Increasing Sleepiness | 1.10 (0.43,2.84) | 2.65 (0.69,10.16) | 1.11 (0.43,2.86) | 3.33 (0.83,13.31) |
| **Total sleep time** |  |  |  |  |
| Stable/improving | 1.00 (ref) | 1.00 (ref) | 1.00 (ref) | 1.00 (ref) |
| Worsening | 0.54 (0.18,1.64) | 0.29 (0.04,2.40) | 0.54 (0.18,1.64) | 0.31 (0.04,2.56) |
| **Sleep efficiency** |  |  |  |  |
| Stable/improving | 1.00 (ref) | 1.00 (ref) | 1.00 (ref) | 1.00 (ref) |
| Worsening | 1.51 (0.47,4.92) | **4.09 (1.13,14.74)** | 1.50 (0.46,4.87) | 3.73 (1.00,13.92) |
| **Wake after sleep onset** |  |  |  |  |
| Stable/improving | 1.00 (ref) | 1.00 (ref) | 1.00 (ref) | 1.00 (ref) |
| Worsening | 0.77 (0.20,3.02) | **6.14 (1.69,22.22)** | 0.76 (0.19,2.99) | **5.76 (1.55,21.48)** |
| **Nap duration** |  |  |  |  |
| Stable/improving | 1.00 (ref) | 1.00 (ref) | 1.00 (ref) | 1.00 (ref) |
| Worsening | 2.14 (0.79,5.79) | 1.45 (0.38,5.56) | 2.17 (0.80,5.92) | 1.69 (0.43,6.61) |
| **Nap frequency** |  |  |  |  |
| Stable/improving | 1.00 (ref) | 1.00 (ref) | 1.00 (ref) | 1.00 (ref) |
| Worsening | 1.29 (0.49,3.38) | 1.07 (0.26,4.45) | 1.29 (0.49,3.39) | 1.06 (0.25,4.59) |
| **Acrophase** |  |  |  |  |
| Stable/later | 1.00 (ref) | 1.00 (ref) | 1.00 (ref) | 1.00 (ref) |
| Earlier | 0.59 (0.18,1.92) | 1.2 (0.30,4.78) | 0.60 (0.18,1.93) | 1.41 (0.34,5.78) |
| **Amplitude** |  |  |  |  |
| Stable/improving | 1.00 (ref) | 1.00 (ref) | 1.00 (ref) | 1.00 (ref) |
| Worsening | 2.36 (0.94,5.88) | 1.52 (0.45,5.11) | 2.38 (0.95,5.94) | 1.74 (0.51,5.92) |
| **Mesor** |  |  |  |  |
| Stable/improving | 1.00 (ref) | 1.00 (ref) | 1.00 (ref) | 1.00 (ref) |
| Worsening | 2.26 (0.82,6.25) | 1.18 (0.28,4.91) | 2.27 (0.82,6.27) | 1.30 (0.31,5.43) |
| **Robustness** |  |  |  |  |
| Stable/improving | 1.00 (ref) | 1.00 (ref) | 1.00 (ref) | 1.00 (ref) |
| Worsening | 0.69 (0.26,1.87) | 1.22 (0.39,3.82) | 0.70 (0.26,1.89) | 1.49 (0.46,4.82) |
| Note: Odds ratios and 95% confidence intervals are reported. Total N=215.  Abbreviations: MCI, mild cognitive impairment; APOE, apolipoprotein E.  *Both models adjusted for age, race, education, body mass index, diabetes, hypertension, myocardial infarction, antidepressant use, and baseline cognition. The APOEε4 status-adjusted model further adjusted for APOEε4 status (at least one APOEε4 allele vs. none). | | | | |
